# Neurogenetic Mechanisms of Risk for ADHD: Examining Associations of Functionally-Annotated Polygenic Scores and Brain Volumes in a Population Cohort

**DOI:** 10.1101/2022.12.12.22283356

**Authors:** Quanfa He, Taylor Keding, Qi Zhang, Jiacheng Miao, Ryan J. Herringa, Qiongshi Lu, Brittany G. Travers, James J. Li

## Abstract

**Background:** ADHD polygenic scores (PGS) are reliably predictive of ADHD outcomes across studies. However, traditional PGS are *statistical* indices of genetic liability – predictive of ADHD but uninformed by *biological* information. The objective of our study was to determine whether a novel, biologically-informed, functionally annotated ADHD PGS can reveal specific brain mechanisms of risk for ADHD.

**Methods:** Data were from the Philadelphia Neurodevelopmental Cohort (*n*=555). Multiple mediation models were tested to examine the indirect effects of ADHD PGS (including one using a functionally annotated approach, i.e., *AnnoPred*) on ADHD inattention (IA) and hyperactivity-impulsivity (HI) symptoms, via gray matter volumes in the cingulate gyrus, angular gyrus, caudate, dorsolateral prefrontal cortex (DLPFC), and inferior temporal lobe. Age-stratified analyses (children, adolescents, and young adults) were also conducted to account for developmental differences.

**Results:** A direct effect was detected between *AnnoPred* PGS and IA symptoms in adolescents only. No indirect effects via brain volumes were detected for either IA or HI symptoms. However, ADHD PGS were consistently associated with the DLPFC.

**Conclusions:** A biologically-informed PGS yielded a more powerful (and developmentally-specific) signal in detecting a direct effect of ADHD PGS on adolescent IA symptoms. However, no indirect effects between ADHD PGS and ADHD symptomology via the brain emerged. ADHD will become increasing predictive as discovery sample sizes climb. Studies that integrate both genetic and neuroimaging information are critical to advancing our understanding of the biological mechanisms underlying ADHD.

---

Attention-deficit/hyperactivity disorder (ADHD) is a neurodevelopmental disorder that affects 3% - 6% of youths and adults worldwide (1,2). Genes explain a substantial proportion of variability in ADHD (3), accounting for an estimated 70 to 80% of its variation (4,5). Genome-wide associations studies (GWAS) identified genetic variants that are associated with ADHD, including several replicated loci across diverse samples (7,8). However, the biological significance of these loci, particularly as they pertain to the structure and/or function of measured brain-based markers, have yet to be examined in human studies.

ADHD is a highly polygenic disorder, in which there are many genes of individual small effects that contribute to its etiology (3,9,10). Polygenic scores (PGS) are widely used in characterizing a person’s polygenic risk for a trait of interest (11). PGS are traditionally computed as the sum of the number of risk alleles at each genetic loci weighted by their effect sizes, which are extracted from summary statistics of a GWAS conducted in an independent sample. In two recent meta-analyses, traditional ADHD PGS reliably predicted ADHD outcomes across samples and populations, explaining as much as 4% of the variance (3,10). By comparison, the most statistically significant genetic variant in the ADHD GWAS itself only accounted for .1% of the variance in ADHD (7). Notably, the predictive power of PGS is expected to grow as GWAS sample sizes for ADHD increase over time (3).

However, enthusiasm around the use of ADHD PGS may need to be tempered by the fact that they are statistically-derived (using GWAS summary statistics) predictors with unknown biological plausibility (3). A functional annotation of the ADHD GWAS showed that genetic variants associated with ADHD were significantly overrepresented in regulatory regions of DNA in cells in the anterior caudate, cingulate gyrus, angular gyrus, dorsolateral prefrontal cortex (DLPFC), and inferior temporal lobe, suggesting that ADHD-associated variants may involve the regulation of gene expression in these five brain regions (7). Furthermore, several of the GWAS-identified ADHD genes (e.g., *SORCS3, DUSP6, SEMA6D*) are involved in neurotransmission, neuronal development, and neuronal plasticity, suggesting the plausibility of brain-based mechanisms underlying ADHD PGS via genetic effects. However, biological annotations of GWAS are not traditionally incorporated in computations of PGS.

To leverage functional annotation information, we employed a novel method known as *AnnoPred*, which upweights genetic variants located in annotation categories that are overrepresented in GWAS (12). Annotation categories included enhancers, promoters, conserved region, coding region, transcription factor binding sites, and cell-type specific epigenomic marks in central nervous system, immune system, cardiovascular system, gastrointestinal system, and skeletal muscles. Upweighted genetic variants are considered more functionally relevant (by effect size) for a trait of interest. In simulated and empirical data, this method demonstrated improved accuracy in disease stratification compared to other PGS methods (12).

In addition to gene enrichment findings from ADHD GWAS (7), other possible gene-brain mechanisms in ADHD come from the broader neuroimaging literature (6,13). Several studies have reported associations between smaller regional volumes in areas such as the caudate, prefrontal cortex, temporal lobe, cingulate cortex, and cerebellum in samples of children and adolescents with ADHD relative to controls (14–17). In particular, the caudate and dorsolateral prefrontal cortex have been extensively implicated in ADHD studies (18–21). The caudate nucleus is involved in motor movement, associative learning, memory, and goal-directed behavior, and was found to be coactive with the DLPFC and anterior cingulate cortex during a range of cognitive and motor tasks (22). The DLPFC is linked to higher level cognitive function such as inhibition, planning, and working memory (23). Significantly smaller caudate nucleus and DLPFC have been documented in youths with ADHD relative to typically developing youths (16,19). Other studies have shown that brain volume differences between ADHD and controls are more dispersed over the brain than previously hypothesized. For example, several studies have reported volumetric reductions in the temporal lobe and cingulate cortex in youths with ADHD relative to controls (15,20,24,25).

Whereas prior studies investigated genetic and brain volume associations with ADHD as separate mechanisms, Alemany and colleagues (26) examined whether brain volume might *mediate* genetic associations (via PGS) of several psychiatric disorders, including schizophrenia, depression, bipolar disorder, ADHD and autism spectrum disorder. This study tested the association between PGS on five psychiatric disorders and volumes of several brain regions of interest (i.e., amygdala-hippocampus complex, caudate, putamen, thalamus, cerebellum, total brain, ventricles, cortical and subcortical gray matter, and total white matter) in a large, population-based sample of 9-11-year-old children (*n=*1,139). ADHD PGS was specifically associated with smaller caudate volume but not the other regions, and the association between ADHD PGS and male-specific attention problems was mediated by caudate volume, in line with the well-replicated association between caudate and ADHD. Their findings provide compelling (albeit, preliminary) evidence that ADHD PGS may reflect specific biological mechanisms in the development of ADHD.

However, one critical limitation of the prior study (26) is that development was not accounted for. ADHD symptom presentations vary depending on the age of the individual (27,28). For instance, whereas inattention (IA) symptoms tend to be relatively stable throughout development, hyperactivity/impulsivity (HI) symptoms tend to decline by adulthood (27,29). Thus, brain-based differences between ADHD and control samples may be age-dependent. In a mega-analysis of 23 cohorts of ADHD cases and cohorts (*n=*3,200), the researchers found that volumetric differences between ADHD cases and controls were larger among children (aged between 4-14) than among adolescents (aged between 15-21) or among adults (aged 22-63) (30). Specifically, they found that accumbens, amygdala, caudate, hippocampus, putamen, and total intracranial volumes were smaller in ADHD children than controls, whereas only the hippocampus was significantly smaller in ADHD cases in adolescents; no brain volume differences were found in ADHD adults compared to controls. Another study compared whole brain volumes and subcortical regional volumes between ADHD cases and controls in a Dutch sample of youths and adults (*n*=672). They reported a reversal in the direction of difference between cases and controls among young adolescents (aged 8-15), older adolescents (aged 16-22) and young adults (aged 22-30), such that caudate and putamen volumes were smaller in ADHD cases than controls among young adolescents, but larger in ADHD cases among young adults (31). The more restricted and younger age range of the adult sample in the latter study (31) likely contributed to the inconsistent findings compared to the first study (30). In light of these inconsistent findings, developmental considerations must be taken into account in genetically-informed studies of neural mechanisms underlying ADHD.

This study focused on children (aged 8-11), adolescents (aged 12-17), and young adults (aged 18-21) from the Philadelphia Neurodevelopmental Cohort (PNC). Although ADHD PGS are frequently examined in clinical samples, they are also predictive of psychiatric traits in population samples as well, as demonstrated in the meta-analysis of ADHD PGS across several studies (3). Examining PGS in population-based cohorts like PNC can critically extend our knowledge about the neurogenetic basis of ADHD in a normative sample. We specifically examined whether brain volumes of regions of interests (i.e., caudate, cingulate gyrus, angular gyrus, DLPFC, and inferior temporal lobe) will mediate the association between PGS and ADHD. PGS was computed in two ways, the first as a “traditional” PGS (11) and the second that leverages functional annotations of GWAS via *AnnoPred* (12). First, we hypothesized that both the traditional and *AnnoPred* PGS will associate with ADHD outcomes (i.e., HI and IA symptoms) across age groups. Second, and based on previously reported age-related brain volume differences between ADHD cases and controls across age groups, we hypothesized that all five brain regions will specifically mediate the path between *AnnoPred* ADHD PGS (i.e., a more biologically sensitive measure of polygenic liability over traditional ADHD PGS) and ADHD outcomes in children and adolescents, but not in adults. We made no hypothesis regarding potential differences in the indirect effects for ADHD IA and HI symptoms given that there have been no strong lines of evidence suggesting differential ADHD PGS associations by ADHD presentation type.

## Method

### Participants

The Philadelphia Neurodevelopmental Cohort (PNC) is a population-based cohort of children, adolescents, and young adults with data collected on psychiatric disorders, medical history, neuroimaging, genetics, and neurocognition. Participants were recruited in the greater Philadelphia, Pennsylvania area between November 2009 to December 2011 (32). Psychiatric disorders were assessed using a semi-structured, computerized clinical interview adapted from the Kiddie Schedule for Affective Disorders and Schizophrenia (K-SADS). The interview was administered to the caregivers or legal guardians (i.e., collaterals) for participants aged 8 to 10 (i.e., children; analytic *n=*137), to participants and collaterals for participants aged 11 to 17 (i.e., adolescent; analytic *n=*297), and the participants themselves if they were between the ages of 18 to 21 (i.e., young adults; analytic *n=*121). To somewhat account for the fact that different age groups utilized different raters, we used data from collaterals for participants between 8 to 17 years of age, and from self-report for those older than 18. For additional details on phenotyping and coding, please refer to He and Li (33). Unfortunately, due to known population stratification effects in ad-mixed samples and the fact that PGS are highly (and problematically) underpowered to predict outcomes in non-European ancestry populations (34,35), we focused the current analyses on PNC individuals who self-reported as European ancestry with genotype, phenotypic and neuroimaging data (after quality control) (full analytic *N=*555).

### Brain imaging and data processing

Neuroimaging details for the PNC sample are provided in Sattherwaite et al. (32), and are briefly summarized here. Data were acquired from a 3T Siemens TIM Trio scanner at the University of Pennsylvania. The structural images used for this study were obtained via a magnetization-prepared 180 degrees radio-frequency pulses and rapid gradient-echo (MPRAGE) sampling sequence (TR = 1810 ms, TE = 3.5 ms, 160 1 mm slices).

Freesurfer (v. 7.1.1) (36) was used to automatically parcellate and segment the five regions of interest (ROIs) used in the study (caudate, cingulate gyrus, angular gyrus, dorsolateral prefrontal cortex, and inferior temporal lobe). These regions were pre-selected based on previous evidence of enrichment for ADHD genetic associations (3,7). The command *recon_all*.,with all default parameters taken, was used with processing steps briefly summarized here; the processing stream performs the following sequential steps: skull stripping, volumetric labeling, intensity normalization, white matter segmentation, surface atlas registration, surface extraction, and gyral labeling. The *Destrieux* atlas (37) was used to extract gray matter volume estimates from ROIs by hemisphere. Bilateral ROI volumes were used as the mediators, per empirical precedent (38).

### ADHD Symptoms

IA and HI ADHD symptoms were assessed from six and three items administered on the adapted K-SADS, respectively. Example items included: “Did you often have trouble paying attention or keeping your mind on your school, work, chores, or other activities that you were doing?”, “Did you often have trouble making plans, doing things that had to be done in a certain kind of order, or that had a lot of different steps?” and “Did you often blurt out answers to other people’s questions before they finished speaking or interrupt people abruptly?” Symptom counts of the two ADHD dimensions were computed by summing the number of endorsed items for each dimension.

### Genotyping and Polygenic scores

#### Genotyping in PNC

All participants were genotyped upon consent using common SNP arrays including Affymetrix Affy60 and Axiom, and Illumina HumanHap550 (v1, v3), Human610-Quad (v1), and HumanOmniExpress. We conducted standard quality control and imputation, excluding SNPs with call rate <95%, minor allele frequency <5%, Hardy-Weinberg equilibrium p<1*10^−4^ (32,39).

#### Discovery GWAS

We used the most recent ADHD GWAS, a case-control meta-analysis that consisted of 55,374 children and adults (20,183 cases and 35,191 controls) from 12 studies of mixed (but predominantly European) ancestries (7). The largest cohort among these 12 studies is a population-based case-control cohort in Denmark (iPSYCH; 14,584 cases and 22,492 controls). The other 11 case-control or trio samples were collected in Europe, Canada, United States and China, and aggregated by the Psychiatric Genomic Consortium (PGC). ADHD case status was determined based on International Classification of Diseases, tenth revision (ICD-10) in iPSYCH, and semi-structured clinical interviews (e.g., Schedule for Affective Disorders and Schizophrenia for School-Age Children, K-SADS) in the other 11 cohorts.

#### Traditional PGS

To construct the traditional PGS, we used PLINK (40) to clump the ADHD GWAS summary statistic and used the European ancestry samples in 1000 Genomes Project Phase III cohort (41) as the linkage disequilibrium (LD) reference panel. We then specified an LD window size of 1000 kb and a r^2^ threshold of 0.1 for clumping. Finally, we generated PGS in the PNC target sample in *PRSice-2* (42) and specified a *p*-value threshold of 1 to include all SNPs.

#### AnnoPred PGS

We constructed annotation-informed PGS for ADHD using *AnnoPred* (12), a Bayesian framework that leverages genomic annotation information to improve polygenic risk prediction. This approach improves SNP effect size estimation by integrating publicly available ADHD GWAS (7) and ADHD heritability enrichment in various functional annotation categories estimated using LD score regression (LDSC) (43). We incorporated 53 baseline annotations in LDSC (43), GenoCanyon annotation quantifying the overall genomic functionality (44), and 66 GenoSkylinePlus cell-type specific annotations (45) to improve the ADHD *AnnoPred* score. We used 1000 Genomes Project European samples (41) as the LD reference and the infinitesimal prior in *AnnoPred* to produce SNP posterior mean effects.

### Analyses

We fit parallel multiple mediation models that included the five brain systems as mediators, ADHD PGS (traditional and *AnnoPred*) as predictors, and ADHD IA and HI symptoms as outcomes. Parallel multiple mediation models were conducted to account for expected correlations among volumes of the five brain regions. Given the number of models and tests conducted, *p*-values were FDR-adjusted using the Benjamin-Hochberg method (46). In age-stratified analyses, we divided the sample by age groups (8-11-year-olds were “children”, 12-17-year-olds were “adolescents”, and 18-21-year-olds were “young adults”) to test age-specific effects of PGS and indirect effects. Good model fit was defined as Comparative Fit Index (>.95) and Root Mean Square Error of Approximation (<.06) (47). Analyses were conducted in R *4*.*1*.*2* using the following packages: *semTools, lavaan, psych, stats*. Age, biological sex, standardized total intracranial volume and the first 10 genetic principal components (to account for population stratification) were included as covariates in all models.

## Results

### Descriptive Statistics

The average age of the PNC sample was 14.36 (*sd*=3.39), with 272 (49.10%) being female. When stratified by age groups (i.e., children, adolescents, and young adults), the average age was 9.672 (*sd=*1.058), 14.790 (*sd*=1.710) and 18.64 (*sd=*0.837), respectively. Furthermore, 46.715% (*n=*64), 46.464% (*n=*138) and 57.851% (*n=*70) were females across these age groups, respectively. Participants had an average of 2.489 (*sd=*3.080), 2.037 (*sd*=2.883) and 2.380 (*sd*=2.675) ADHD symptoms, respectively. More details of the sample are shown in Table 1, including standardized (in the full sample) regional brain volumes and total intracranial volume.

**Table 1.** Descriptive statistics

|  | Full | Children | Adolescents | Young Adults |
| --- | --- | --- | --- | --- |
| *n* | 555 | 137 | 297 | 121 |
| Age (*sd*) | 14.360 (3.394) | 9.672 (1.058) | 14.790 (1.710) | 18.640 (.837) |
| Biological sex (# female/percentage) | 272 (49.100%) | 64 (46.715%) | 138 (46.464%) | 70 (57.851%) |
| ADHD total symptoms (*sd*) | 2.223 (2.891) | 2.489 (3.080) | 2.037 (2.883) | 2.380 (2.675) |
| ADHD inattention symptoms (*sd*) | 1.634 (2.173) | 1.759 (2.235) | 1.562 (2.249) | 1.669 (1.908) |
| ADHD hyperactivity/impulsivity symptoms (*sd*) | .589 (.967) | .729 (1.068) | .475 (.893) | .711 (.995) |
| Brain regions (volume) |  |  |  |  |
| Angular gyrus, standardized (*sd*) | -.010 (.867) | .458 (.872) | -.049 (.803) | -.044 (.754) |
| Caudate, standardized (*sd*) | -.011 (.975) | .302 (.935) | -.068 (.987) | -.227 (.909) |
| Cingulate gyrus, standardized (*sd*) | -.013 (.988) | .469 (.874) | -.044 (.984) | -.485 (.872) |
| DLPFC, standardized (*sd*) | -.015 (.983) | .547 (.934) | -.073 (.934) | -.511 (.906) |
| Inferior temporal lobe, standardized (*sd*) | -.008 (.996) | .328 (.890) | .022 (.947) | -.460 (1.062) |
| Total intracranial volume, mm<sup>3</sup> (*sd*) | 1413671<br>(143764.2) | 1417708<br>(114153.5) | 1421687<br>(146137.3) | 1389422<br>(164925.8) |

### Traditional ADHD PGS – Full Sample

Multiple mediation models for IA and HI symptoms fit the data well (CFI=.995, RMSEA=.096 for both models). Total and direct effects were not significant for either HI or IA symptoms with traditional ADHD PGS as the predictor (Figure 1; *FDR*-corrected *p* >.05). No indirect effects emerged with traditional ADHD PGS on HI or IA symptoms via the angular gyrus, caudate, cingulate gyrus, DLPFC, and inferior temporal lobe. Traditional ADHD PGS was associated with DLPFC (*b=*-.084, *se=*.028, *FDR*-corrected *p*=.006), but not with the other four brain regions. In addition, the paths from cingulate gyrus (but not the other brain regions) to HI and IA symptoms were significant (*b=*.179, *se=*.064, *FDR*-corrected *p*=.010; *b=*.295, *se=*.144, *FDR*-corrected *p*=.041).

**Figure 1.**
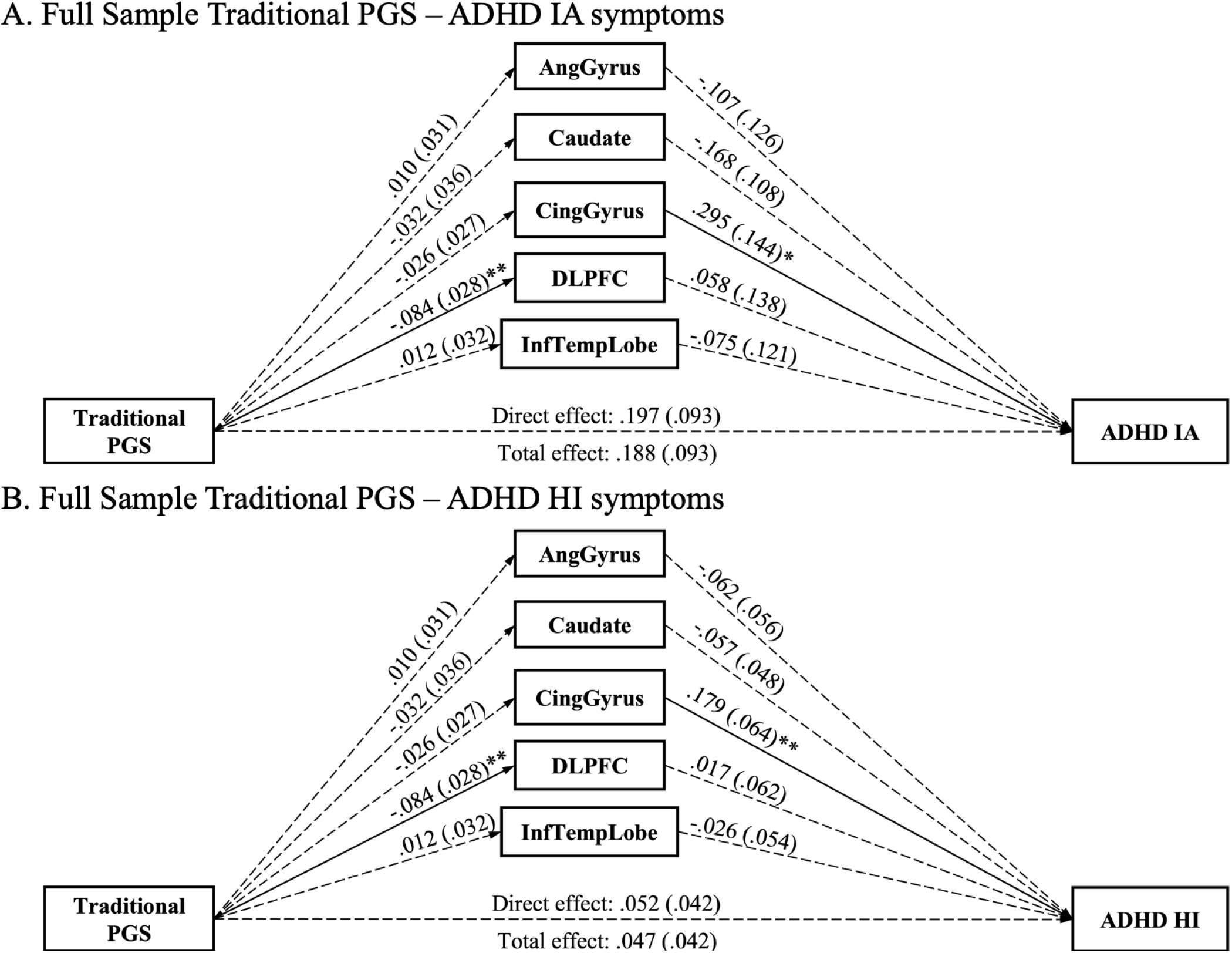
Multiple mediation paths between traditional ADHD PGS and ADHD symptoms via five brain regions in the full sample of youths and young adults in PNC.

### Traditional ADHD PGS – Age-Stratified Models

The same models were tested separately in each of the three age groups to investigate the possibility of developmental differences of traditional ADHD PGS and their effects through regional brain volumes on IA and HI symptoms. In children (*n*=137), the models fit well for IA and HI symptoms (CFI=.999, RMSEA=.042 for both models). No total, direct, or indirect effects via the five brain regions were detected (Figure 2). Traditional ADHD PGS was not associated with any of the five regional volumes. However, the angular gyrus volume (but not other brain regions) was significantly associated with IA symptoms (*b=*-.561, *se=*.232, *FDR*-corrected *p*=.032).

**Figure 2.**
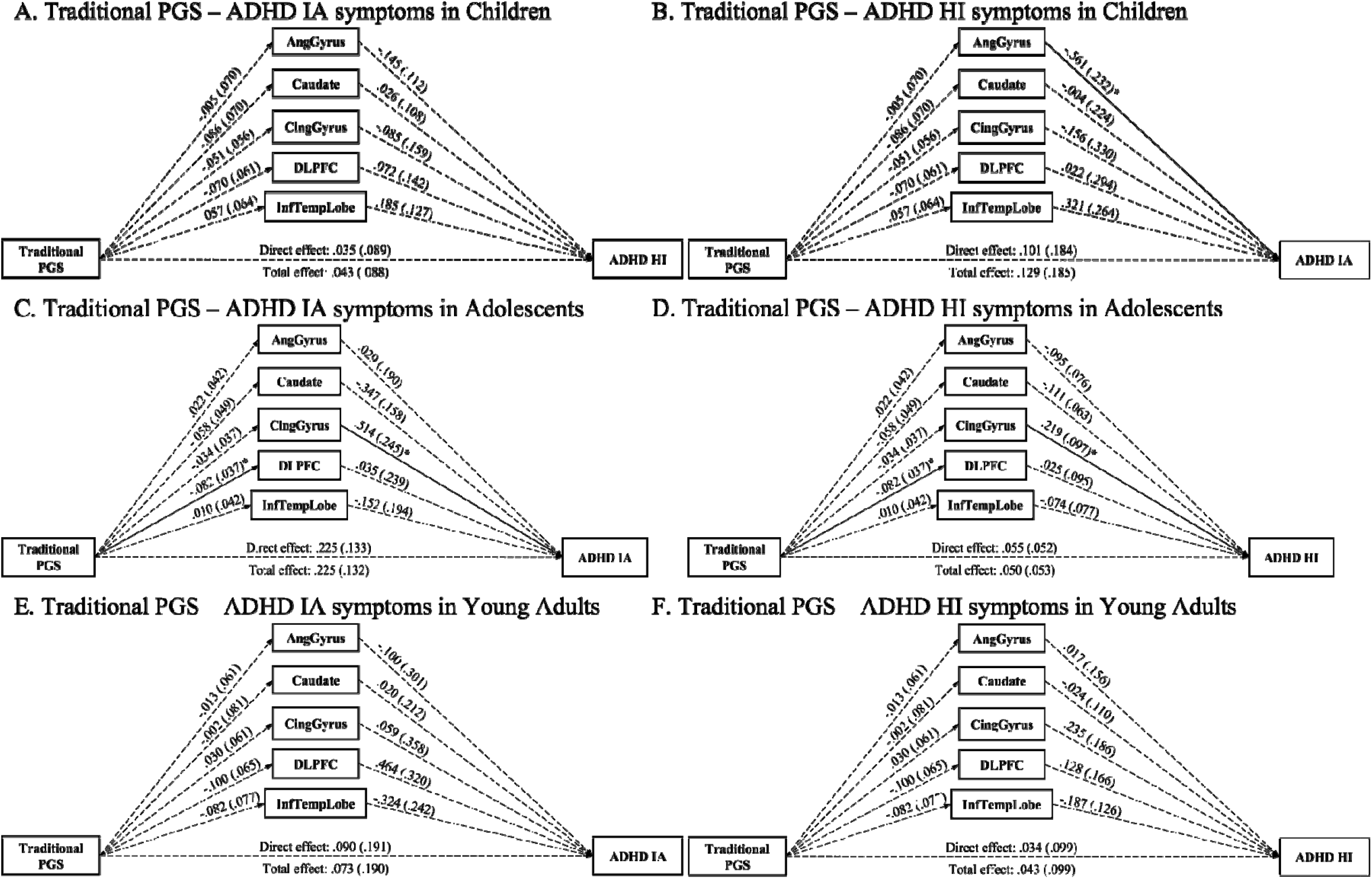
Multiple mediation paths between traditional ADHD PGS and ADHD symptoms via brain regions in children, adolescents, and adults in PNC

Similarly, no total, direct, or indirect effects were detected in adolescents *(n=*297) (Figure 2). The models for IA and HI symptoms fit acceptably (CFI=.992, RMSEA=.118 for both models). The path from traditional PGS to DLPFC (but not other regions) was significant (*b=*-.082, *se=*.037, *FDR*-corrected *p*=.050). The cingulate gyrus (but not other brain regions) was associated with HI and IA symptoms (*b=*.219, *se=*.097, *FDR*-corrected *p*=.036; *b=*.514, *se=*.245, FDR-corrected *p*=.036, respectively).

No total, direct or indirect, PGS-brain nor brain-ADHD pathways were detected in young adults (see Figure 2). The models fit well for both IA and HI symptoms (CFI=.998, RMSEA=.032 for both models).

### AnnoPred ADHD PGS – Full Sample

The models for IA and HI symptoms fit well (CFI=.995, RMSEA=.098 for both models). There were no total effects, total indirect, or direct effects of *AnnoPred* ADHD PGS on HI or IA symptoms (Figure 3). However, the path from *AnnoPred* ADHD PGS to the DLPFC (but not other brain regions) was significant (*b=*-.063, *se=*.029, *FDR*-corrected *p*=.030). Additionally, the paths from the cingulate gyrus (but not other regions) to HI and IA symptoms were both significant (*b=*.181, *se=*.064, FDR-corrected *p*=.010; *b=*.301, *se=*.144, *FDR*-corrected *p*=.041, respectively).

**Figure 3.**
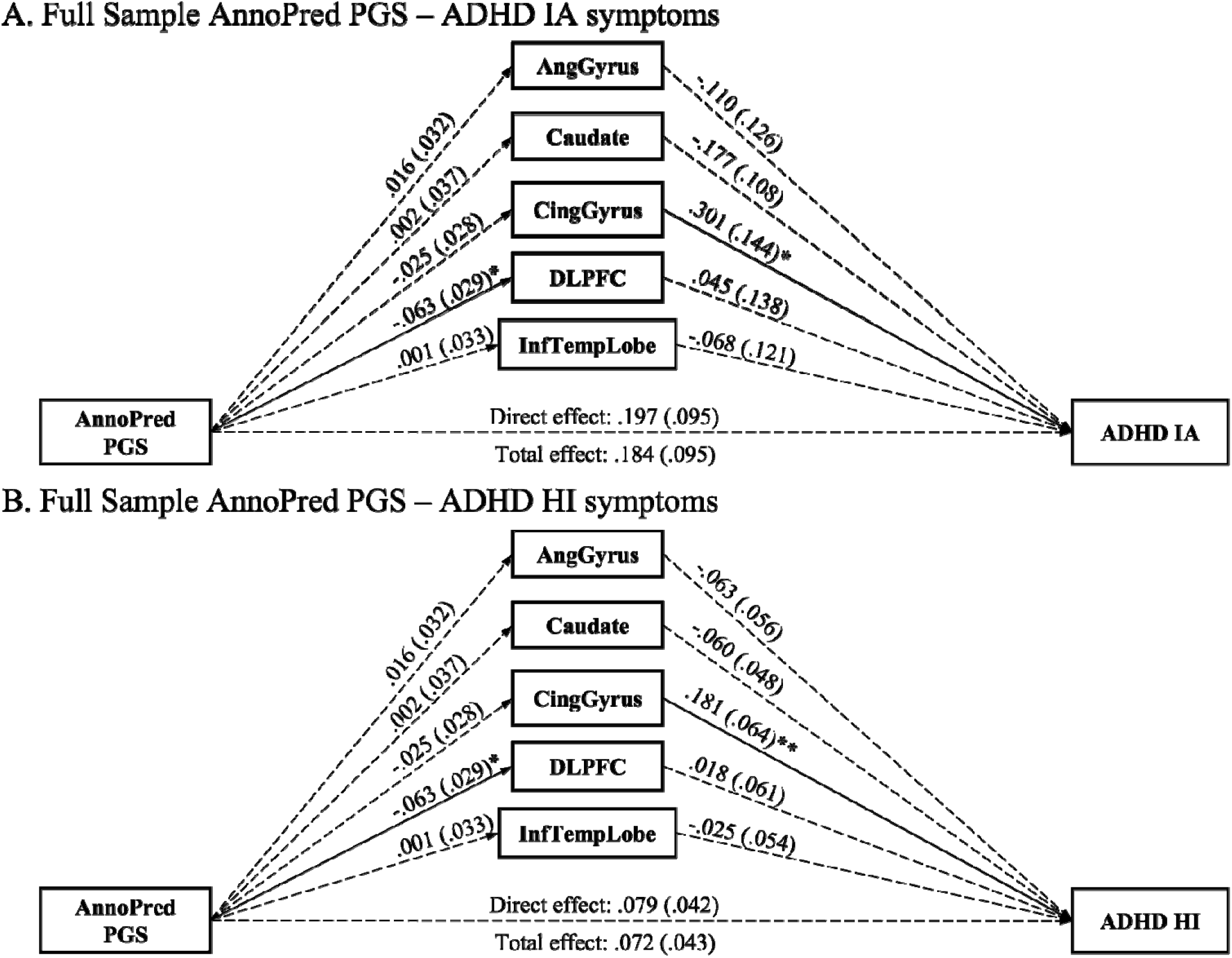
Multiple mediation paths between AnnoPred ADHD PGS and ADHD symptoms via brain regions in the full sample of children, adolescents, and young adults in PNC

### AnnoPred ADHD PGS – Age-Stratified Models

In children, the models fit well for IA and HI symptoms (CFI=.998, RMSEA=.051 for both models). No total or direct effects of *AnnoPred* ADHD PGS on HI and IA symptoms or indirect effects via the five brain regions were detected (Figure 4). *AnnoPred* ADHD PGS was not associated with any of the five brain regions. The angular gyrus (but not other brain regions) volume was associated with IA symptoms (*b=*-.594, *se=*.233, *FDR*-corrected *p*=.032). No regional volume was associated with HI symptoms.

**Figure 4.**
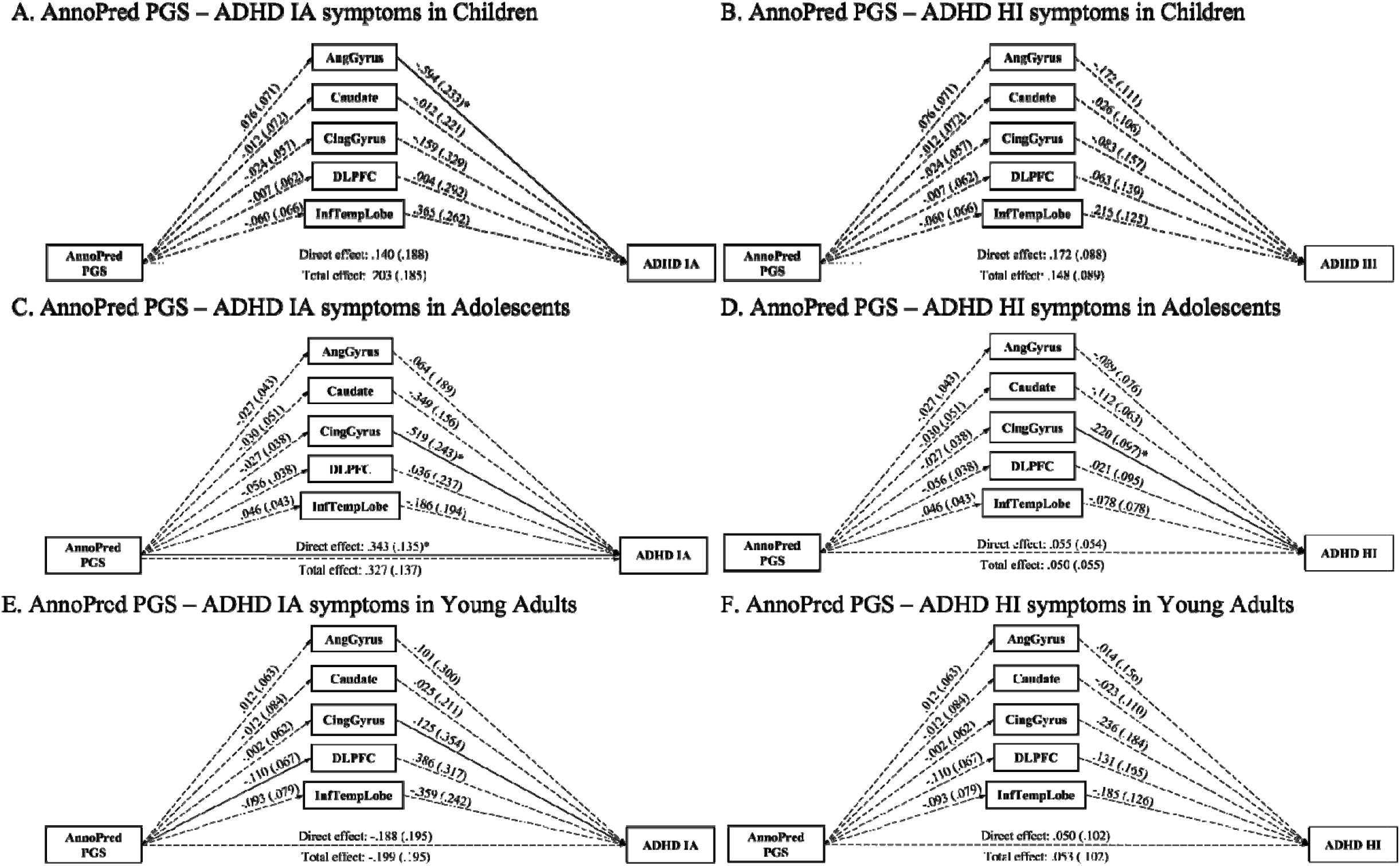
Multiple mediation paths between AnnoPred ADHD PGS and ADHD symptoms via brain regions in children, adolescents, and young adults in PNC

In adolescents, the models fit acceptably for IA and HI symptoms (CFI=.992, RMSEA=.118 for both models). There was a direct effect from *AnnoPred* ADHD PGS to IA symptoms (*b=*.343, *se=*.135, FDR-corrected *p*=.044), but not for HI symptoms. No total or indirect effects emerged for IA and HI symptoms via brain volumes (Figure 4). *AnnoPred* ADHD PGS was not associated with any of the five regional volumes. However, the paths from cingulate gyrus to HI symptoms and IA symptoms were significant (*b=*.220, *se=*.097, FDR-corrected *p*=.036; *b=*.519, *se=*.243, *FDR*-corrected *p*=.036).

Finally, in young adults, the models fit well for IA and HI symptoms (CFI=.999, RMSEA=.032 for both models). No total, direct, or indirect effects emerged for PGS-brain or brain-ADHD pathways to either IA or HI symptoms (Figure 4).

## Discussion

The primary objective of this study was to investigate whether the effects of ADHD PGS, computed both traditionally and annotated functionally, were mediated by volumes of five brain regions – the caudate, cingulate gyrus, angular gyrus, DLPFC, and inferior temporal lobe – in a population-based dataset of children, adolescents, and young adults. We also examined age-stratified effects of the potential mediation pathways between PGS and ADHD symptoms. With respect to our first hypothesis, we found that traditionally computed ADHD PGS was not associated with either IA or HI symptoms, regardless of age group. However, *AnnoPred* ADHD PGS associated with IA symptoms, but only in adolescents. With respect to our second hypothesis, we found no indirect effects between either the traditional or the *AnnoPred* ADHD PGS via the five brain regions on IA or HI symptoms. No indirect effects between ADHD PGS and IA and HI symptoms via brain volumes were observed in our full sample analyses as well as in our age-stratified models.

First, we found that the *AnnoPred* ADHD PGS was associated with IA (but not HI) symptoms for adolescents specifically. Functionally annotated ADHD PGS may be more developmentally sensitive than traditional ADHD PGS, which have previously been shown to be less discriminative between various age groups (3,10). One possibility is that there may be developmentally distinct genetic influences for ADHD that depends on the age and developmental stage of the individual (1,48–51). This possibility is supported by recent evidence from another ADHD GWAS, which found distinct genetic architectures underlying childhood ADHD, persistent ADHD, and those with adult-onset ADHD (52). This finding supports the possibility that the genetic liability for ADHD may be developmentally specific. Developmental specificity may also help to explain why *AnnoPred* ADHD PGS was associated with IA but not HI symptoms, given the relative stability of IA symptomology over time in individuals (27).

Our findings did not support our second hypothesis regarding indirect effects between *AnnoPred* ADHD PGS and IA and HI symptoms via brain volumes. The lack of significant PGS-brain and brain-ADHD pathways could be due to the non-clinical nature of PNC, which featured a relatively low prevalence of ADHD diagnoses and symptomology. It is possible that the relatively low levels of variability in ADHD symptomology, even for a population-based sample like PNC, may have limited the power to detect effects via ADHD PGS. In addition, most structural MRI studies in ADHD and the ADHD GWAS were conducted in case-control samples of youths or adults (53). The brain regions we focused on in our study were informed by these ADHD case-control studies, which may not have been generalizable to large population-based studies (54). Future studies should examine the possibility that neurogenetic mechanisms of ADHD may differ between clinical and the general population samples (55).

However, we did detect a negative association between ADHD PGS (both traditional and *AnnoPred*) and DLPFC volumes in our study. Although the DLPFC was not associated with ADHD symptoms in our study (perhaps due to the limited variability in ADHD symptoms), prior studies have found that the DLPFC was the most enriched region with respect to ADHD-associated genetic variants relative to the other brain regions we tested (7,19). In our age-stratified analyses, we also observed a negative association between ADHD PGS and DLPFC in adolescents specifically, which might be attributable to an increased heritability of DLPFC volume from childhood to adolescence (56,57). Given that our study was cross-sectional, future studies should explore changes in the heritability of brain measures in relation to the development of ADHD.

We also found several brain volume-ADHD associations that were somewhat inconsistent with the prior neuroimaging literature. We found that *increased* cingulate gyrus volume was associated with greater HI and IA symptoms in the full sample and in adolescents specifically. This was inconsistent from prior literature which found *reduced* volumes in the cingulate cortex in youths and adults with ADHD relative to controls (15,53,58). The cingulate gyrus is part of the cingulate cortex that is known for its role in controlled cognition, response inhibition, novelty detection and motivation (58). We posit two possible explanations for our findings. First, these previous studies regressed volumes of each region of interests separately on ADHD status, along with a set of covariates (e.g., biological sex, age, total intracranial volume). Our study examined the cingulate simultaneously with several other brain regions within a multiple mediator framework (in addition to accounting for standard covariates), thereby accounting for the high degree of covariation between these brain regions. Additionally, these previous studies were conducted in exclusively clinical (or case-control) samples. The cingulate cortex (along with other previously implicated brain regions for ADHD) may associate with natural variations in ADHD symptoms differently in population-based samples that feature lesser severity and greater heterogeneity of symptomologies.

Several limitations in our study should be noted. First, our measure of ADHD in young adults was based on retrospective self-report, whereas ADHD in children and adolescents was based on parent-report. Reporter bias and discrepancies (59) may have affected the generalizability/comparability of our findings between children/adolescents and young adults in our study. Relatedly, self-reported measures of ADHD could be related to lower heritability estimates relative to ADHD reported by other informants (60). Hence, it is possible that the lack of an ADHD PGS direct effect in young adults may be due to this phenomenon. Second, medication status of the sample was unknown, thus preventing us from testing whether our findings are robust against the effect of medication on both ADHD symptoms and on cortical maturation. Third, ADHD HI symptoms were measured by only three items, thus limiting the variability of HI symptoms in the sample. It is possible that the relative lack of variance in HI symptoms can partially explain the developmental specificity of *AnnoPred* PGS and IA symptoms in adolescents, but not HI symptoms. Fourth, we used cortical and subcortical volumes in our study, in line with empirical precedents (38). However, other brain structural measures (e.g., cortical thickness, surface area, fractional anisotropy) as well as functional measures (e.g., regional and network activation) have been studied in relation to ADHD but were not tested in the current study (15,53,54,61). Future studies may consider incorporating other brain measures to test genetic mechanisms of ADHD. Finally, PNC is a cross-sectional study, which precluded our ability to examine whether (or how) PGS affect both brain structures and ADHD symptomology over time (e.g., via longitudinal modeling).

To conclude, we found a developmentally-specific direct effect of novel *AnnoPred* ADHD PGS on IA symptoms in a population-based sample. Our biologically-informed PGS yielded a more powerful and developmentally-specific signal in detecting this direct effect relative to the traditional (i.e., statistically-driven) approach to PGS. However, no indirect effects between ADHD PGS (measured traditionally and *AnnoPred*) and ADHD symptomology via brain regions emerged in our analysis. Of note, although ADHD PGS are robust predictors of ADHD, they only explain about 4% of the variance in ADHD, and even less variance in population-based cohorts (3). As GWAS sample sizes continue to climb, ADHD PGS will become more predictive over time. There will be a need for more studies that integrate both genetic and neuroimaging information to advance our understanding of the biological mechanisms underlying ADHD.

## Data Availability

The data used in this study are publicly available online in dbGaP (https://www.ncbi.nlm.nih.gov/gap/).

https://osf.io/whjuq

## Acknowledgements

Not applicable

## Abbreviations

ADHD: attention deficit hyperactivity disorder
CFI: Comparative Fit Index
DLPFC: dorsolateral prefrontal cortex
GWAS: genome-wide association study
HI: hyperactivity-impulsivity
IA: inattention
PGS: polygenic score
RMSEA: Root Mean Square Error of Approximation

